# Prevalence and Factors Associated with Illicit Drug and High-Risk Alcohol Use among Adolescents Living in Urban Slums of Kampala, Uganda

**DOI:** 10.1101/2023.07.20.23292973

**Authors:** Hellen Kalungi, Onesmus Kamacooko, Jane Frances Lunkuse, Joy Namutebi, Rose Naluwooza, Matt A. Price, Eugene Ruzagira, Yunia Mayanja

## Abstract

**Background:** Illicit drug and high risk alcohol use among adolescents leads to poor health outcomes. We enrolled adolescents from urban slums in Kampala, Uganda, to assess baseline prevalence, and factors associated with illicit drug and high-risk alcohol consumption.

**Methods:** We conducted a cross-sectional study using data collected in a cohort that enrolled 14-19-year-old male and female participants from 25-March-2019 to 30-March 2020. Data was collected on social-demographics, sexual behavior and reproductive health using interviewer administered questionnaires. The main outcomes were illicit drug use and high-risk alcohol use. Data on alcohol use was collected using the Alcohol Use Disorder Identification Test (AUDIT); results were dichotomized. Factors associated with each outcome were analyzed using multivariable logistic regression.

**Results:** We enrolled 490 participants (60.6% female) with median age 18 (IQR 17-18) years, 91.0% had less than secondary education, 48.4% had their sexual debut before 15years, 47.1% reported paid sex in the past 3 months and 24.7% had a sexually transmitted infection (chlamydia, gonorrhea and/ or active syphilis) at enrolment.

The prevalence of illicit drug use was 34.9% while 16.1% were screened as high-risk alcohol users. Illicit drug use was associated with being male (aOR 9.62; 95% CI 5.74-16.11), being married (aOR 2.24; 95%CI 1.07-4.68) and having ≥10 paying sexual partners in the past 3 months (aOR 3.13; 95%CI 1.40-6.98). High risk alcohol use was associated with reporting sex work as the main job (aOR 3.19; 95%CI 1.02-9.94) and having experienced physical (aOR 1.96 95%CI 1.01-3.81) or emotional violence (aOR 2.08; 95%CI 1.14-3.82) from sexual partners.

**Conclusion:** Illicit drug and high-risk alcohol use are prevalent among adolescents involved in high risk sexual behavior and living in urban slums of Kampala. Comprehensive interventions that target substance use among this group of young people are needed and should include measures against intimate partner violence.

## 1. Introduction

According to the 2021 United Nations Office on Drugs and Crime (UNODC) report, the global burden of illicit drug and alcohol use is estimated at 275 million people, an increase of 21.7% from 226 million people in 2010 (1). In 2015 illicit drug use cost the human population tens of millions of disability adjusted life years (DALYs). Europeans suffered proportionately more but in absolute terms the mortality rate was greatest in low and middle income countries (LMICs) where data availability was more limited due to underestimation of illicit drug use (2). DALY rates of illicit drug use are estimated to be 2.5 times higher in Sub-Saharan Africa (SSA) than other regions (3). Alcohol and illicit drug use are a major threat to education, health and the economy (1, 2, 4–6), contributing to 5.1% morbidity and 5.3% mortality worldwide. In 2015, 450,000 people died as a result of illicit drug use; 37.3% of deaths were a direct result of drug use disorders (4, 7). UNODC report that according to the demographic surveys it is projected that by 2030, the number of people using illicit drugs will have risen by 11% globally and 40% in SSA due to its rapidly growing and young population (1). The predictors of illicit drug use among adolescent’s world over include male gender, peer pressure, low education, psychological factors and increasing poverty levels (8–10). Illicit drug and alcohol use contribute to crime, disease, physical and mental incapacitation (1, 5, 11) in both adolescents and adults, however the impact on adolescents with developing minds and bodies may be more profound.

Adolescents are defined by the World Health Organization (WHO) as individuals 10-19 year (12). The global adolescent population is estimated at 1.2 billion, representing one fifth of the global population (3). Surveys among the general population show that the extent of drug use among young people remains higher than among older people. Most research suggests that early (12-14 years) to mid (15-17 years) adolescence is a critical risk period for the initiation of substance use, and that use of substances like alcohol and illicit drugs may peak in late adolescence (18-19 years) and among young people aged 20-24 years (7). Illicit drug and alcohol use increase with age (11, 13) and health behaviors adopted during adolescence such as use of substances have implications that can persist throughout the life course (14). The WHO 2018 global status report on alcohol and health estimates current alcohol use among 15-17 year-olds in SSA at 21.4% (4). Furthermore, a systematic review of studies done in SSA found that the overall prevalence of any substance use among adolescents was 41.6% (15). Reports show that the risk of illicit drug and alcohol use among adolescents in SSA is increased by key drivers such as: transactional sex, availability of disposable income, poverty, gender inequalities and poor work/ living environments (16, 17). Negative effects of illicit drug and alcohol use on adolescents are associated with increased risky behaviors that increase the likelihood of HIV infection, and addictive disorders that commonly result in mental illness (11, 16). There are various family, economic and social factors that predispose adolescents to illicit drug and alcohol use in SSA e.g., prolonged separation from family, unstable housing, availability of drugs in the environment and feelings of loneliness (10, 11, 13). Although a number of interventions have been suggested to reduce illicit drug use in SSA, the evidence of benefit for them is weak (18, 19) and despite the proven dangers of illicit drug use, it persists, and in some contexts increases even with interventions (1). For example, the percentage of adolescents who perceive cannabis as harmful has dropped by 40% despite the evidence linking regular use to health problems particularly in young people and despite the positive correlation between potency and harm (1).

In Uganda, the burden of illicit drug and alcohol use is high among school-going individuals, fisher folk, slum-dwelling adolescents and young women involved in high risk sexual behaviors (9, 10, 17, 20–22) yet this population remains under studied (17). A study done in northern and central Uganda found that two thirds of school going adolescents had ever used drugs (21). Previous studies done in Kampala the capital city and fishing communities in Uganda show that illicit drug and alcohol use among adolescents are associated with sexual risk behaviors such as high numbers of paying sexual partners and inconsistent condom use (10, 17) Unregulated marketing and availability of cheap drugs, alcohol and alcohol branded merchandise to adolescents has increased access to these substances (9, 10, 17, 20). However, few studies have described illicit drug and high-risk alcohol use among adolescents and data are limited among adolescents living in urban slums where use of these substances may be more prevalent.

In this study among male and female adolescents from urban slums in Kampala, we assessed the prevalence and baseline factors associated with illicit drug and high-risk alcohol use.

## 2. Methodology

### 2.1. Study design and setting

We conducted a cross sectional study using data collected in a cohort that recruited adolescents. The main aim of the adolescent cohort was to assess *“**F**easibility of **E**nrolling and **R**etaining **A**dolescents **at R**isk of HIV infection”* (FERDAR). Participants were recruited at the Good Health for Women Project (GHWP) clinic which was established in a peri-urban community in southern Kampala in 2008 to study the epidemiology of HIV and sexually transmitted infections (STIs) and to implement HIV/STI prevention among high-risk women with a focus on female sex workers. The GHWP clinic offered comprehensive HIV prevention, care and treatment services to the women, their children below 5 years and regular male partners.

### 2.2. Study population and recruitment

Recruitment for the FERDAR cohort was done from 25-March-2019 to 30-March-2020 through a project field worker who mobilized 14-19-year-old female and male adolescents from six urban slums in and around Kampala. Participants <18 years met criteria as being mature or emancipated minors as per guidelines of the Uganda National Council for Science and Technology (UNCST) (23). These communities were characterized by sex work, alcohol selling venues and illicit drug consumption. Community awareness sessions were held to inform communities and adolescents about the research activities of the project. Adolescents were initially identified by project field workers with support from community mobilisers and later invited to the GHWP clinic for screening and enrolment. Participants enrolled in the study also mobilized peers using a snowball approach.

#### 2.3. Eligibility criteria

Adolescents who were included in the study were 14-19-years (consenting adolescents 18-19 years, emancipated and/ or mature minors if <18 years) as per the National guidelines (23), sexually active in the last three months, and willing to return to the clinic every 3 months for study procedures.

#### 2.4. Enrolment

Enrolment procedures included HIV counselling and testing), participants who tested HIV positive were enrolled in the study and linked to test and treat program at GHWP for anti-retroviral therapy, screening and treatment for sexually transmitted infections (STIs), a genital swab for STI testing (chlamydia, gonorrhea) and a syphilis sample was collected. Participants found positive with STIs were given treatment, and their partners were invited for testing and treatment. Contraceptive refills and pregnancy tests done for female adolescents and those found pregnant were enrolled and linked to ante-natal care services. We collected data on socio-demographic, sexual behavior and reproductive health variables, illicit drug use in the past three months before enrolment using the social-demographic risk behavior questionnaire and recent alcohol use using AUDIT questionnaire. Illicit drug and high-risk alcohol use counselling was offered to enrolled participants.

### 2.5. Counselling for high-risk alcohol and illicit drug use

Simple advice on reduction of low risk and hazardous drinking (1-7 and 8-15 AUDIT scores, respectively) was given, brief counseling and continued monitoring was provided for high risk/harmful alcohol use (16-19 AUDIT scores), diagnostic evaluation for alcohol dependence (≥20 AUDIT scores) and referral was recommended as in the WHO AUDIT tool guidelines for use in primary care, second edition (24)

### 2.6. Laboratory Procedures

HIV testing was done on serum using the Ministry of Health (MoH) algorithm (Determine, Statpak and SD Bioline) where positive Determine was confirmed by Statpak and SD Bioline was used as a tie-breaker. Serum syphilis testing used the Rapid Plasma Reagin (RPR) for screening and treponema pallidum particle agglutination assay (TPPA) as confirmatory test. A genital swab was taken for STI testing (chlamydia, gonorrhea) using Roche Cobas x4800 Real time polymerase chain reaction. Pregnancy test for females was done on urine, Human Chorionic Gonadotrophin Hormone using pal urine dip stick strip.

### 2.7. Data Collection

Trained research assistants collected data using interviewer administered questionnaires that were translated to the local language of the area (Luganda). Both English and Luganda version of the questionnaires were available depending on the participant’s language of choice.

### 2.8. Study variables

#### 2.8.1. Dependent (outcome) variables included

- Illicit drug use in the 3 months before enrolment. Data was collected as a binary variable (Yes/ No);
- High-risk alcohol consumption at enrolment, a binary variable (Yes/ No). Alcohol use was collected as a numerical variable (Baseline AUDIT score) and then categorized into high risk (AUDIT score ≥16) vs. low to moderate risk (0–15)

#### 2.8.2. Independent variables included

Social demographic variables included participant current age, age at first sexual intercourse, average monthly income, marital status, gender, level of education, sexual and reproductive health variables; the latter included total number of sexual partners in the 3 months before enrolment, number of biological children, number of abortions and miscarriages ever experienced, experience of intimate partner violence (IPV) from sexual partners in the past 3 months (yes/ no), engaging in paid sex in the past 3 months (yes/ no), condom use with partners at the last sexual encounter (yes/ no), type of sexual partner (regular partner living together, regular partners not living together, paying partners, casual acquaintance or any other partners), and contraceptive use for females (yes/ no).

### 2.9. Statistical Analysis

Data were double entered in Open Clinica, cleaned and exported to Stata17.0 (Stata Corp, College Station, TX, USA) for further management and analysis. The participants’ characteristics were summarized in descriptive terms such as mean, median, standard deviations or percentage, as appropriate. The proportions of illicit drug use and high-risk alcohol use at baseline were calculated as proportions of the total number enrolled.

Factors associated with illicit drug use in the past 3 months and high-risk alcohol use at baseline were determined for each outcome and each independent variable using logistic regression. Bivariate/unadjusted logistic regression analysis was conducted and variables with p-value < 0.15 were considered for the adjusted, multivariable analysis. Multivariable logistic regression was used to identify the factors that are independently associated with illicit drugs and high-risk alcohol use. Each outcome was considered separately, with its own model. In the multivariable analysis, variables were kept in the model if removing them significantly affected model fit. Variables that had a P-value <0.05 were considered significantly associated with illicit drug and high-risk alcohol use. In both models, we considered age at enrolment and sex as priori confounders that were included in the multivariable models regardless of its unadjusted p-values.

### 2.10. Ethical considerations

The study was reviewed and approved by the Uganda National council for science and technology (HS 2493) and the Uganda Virus Research Institute Research Ethics committee (GC/127/18/09/658). Written Informed Consent was obtained from every participant.

## 3. Results

### 3.1. Baseline characteristics of enrolled participants

We included 490 participants in our analysis; 297 (60.6%) females and 193 males. The median age of participants was 18 (IQR 17-18) years, 50.8% were ≥18years, 91.0% had less than secondary education and 97.3% were out of school at the time of enrollment. Of 237 (48.4%) adolescents who had their sexual debut before 15years, 56.5% were female vs 43.5% males. Participants who had any STI (chlamydia, gonorrhea and/or active syphilis) at enrollment were 24.7%, of whom 84.7% were female. Of 231 (47.1%) who reported paid sex in the past 3 months, 94.4% were female. Among the 219 females who reported paid sex in the past 3 months, 75.0% had less than secondary education. Adolescents reporting more than one sexual partner were 332 (67.8%) of whom 62.9% were female. Female adolescents using contraception were 59.2%. Of 154 (31.4%) who had biological children 90.9% were females, of whom 10.0% had ≥2 biological children (Table 1).

**Table 1:**
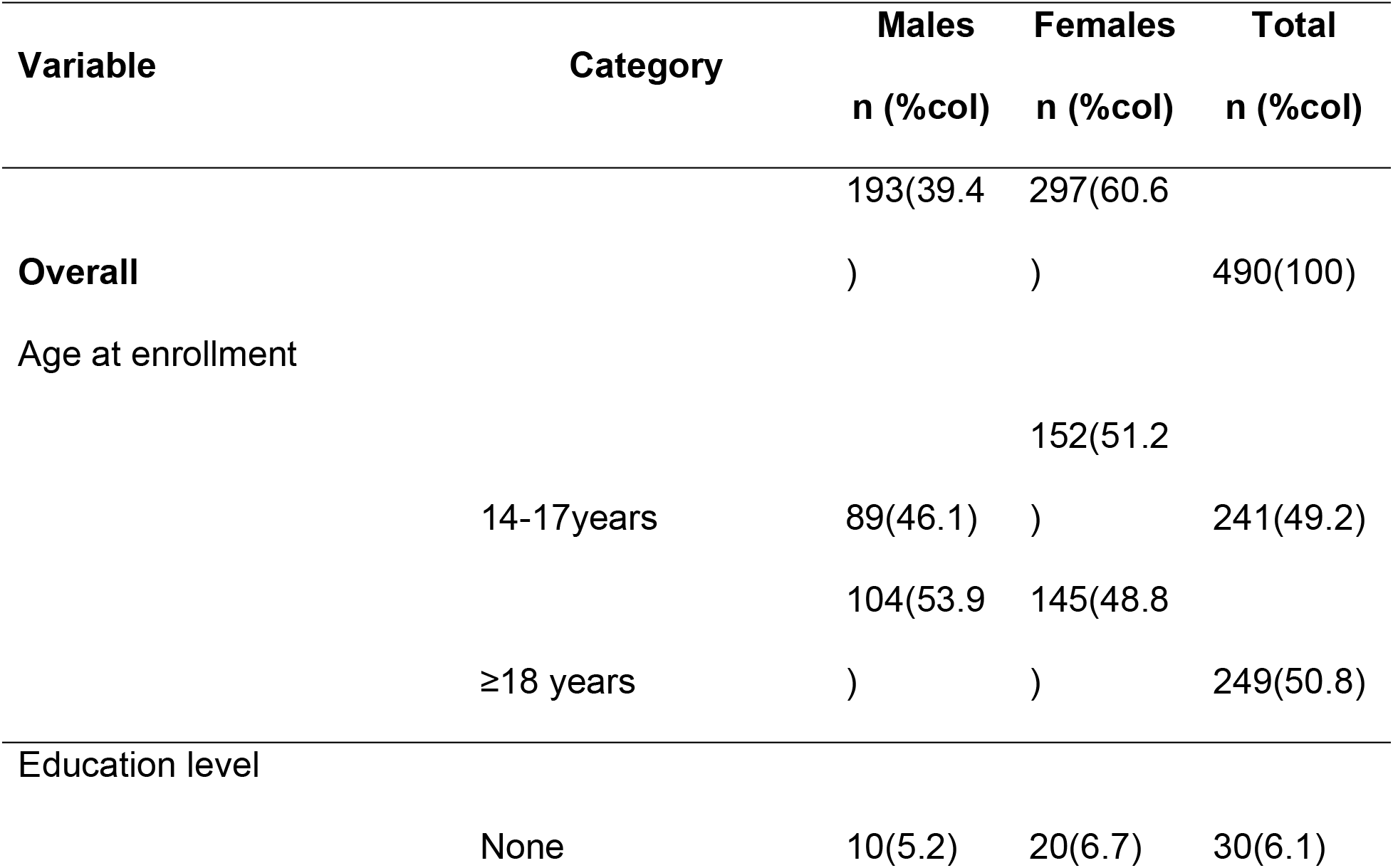
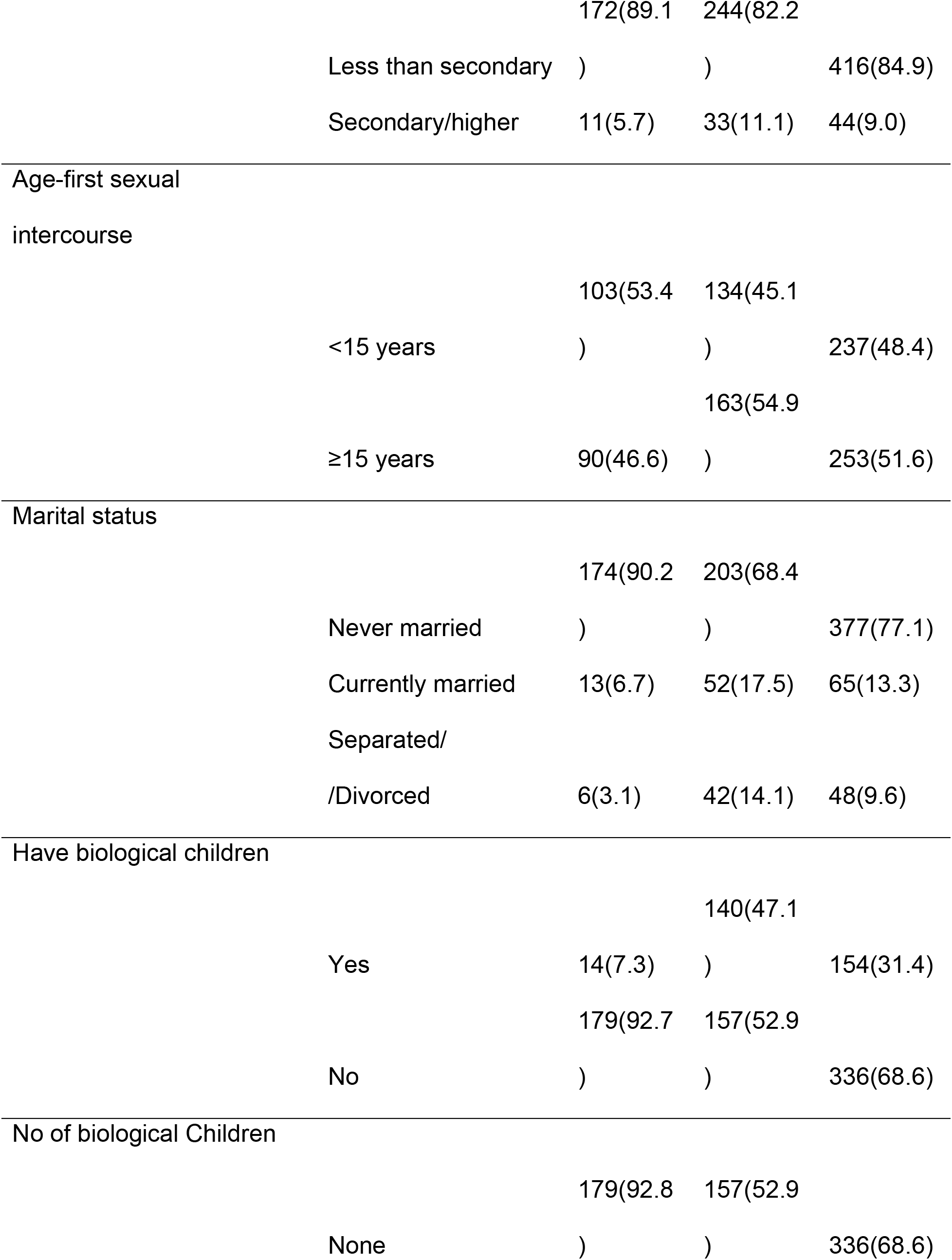
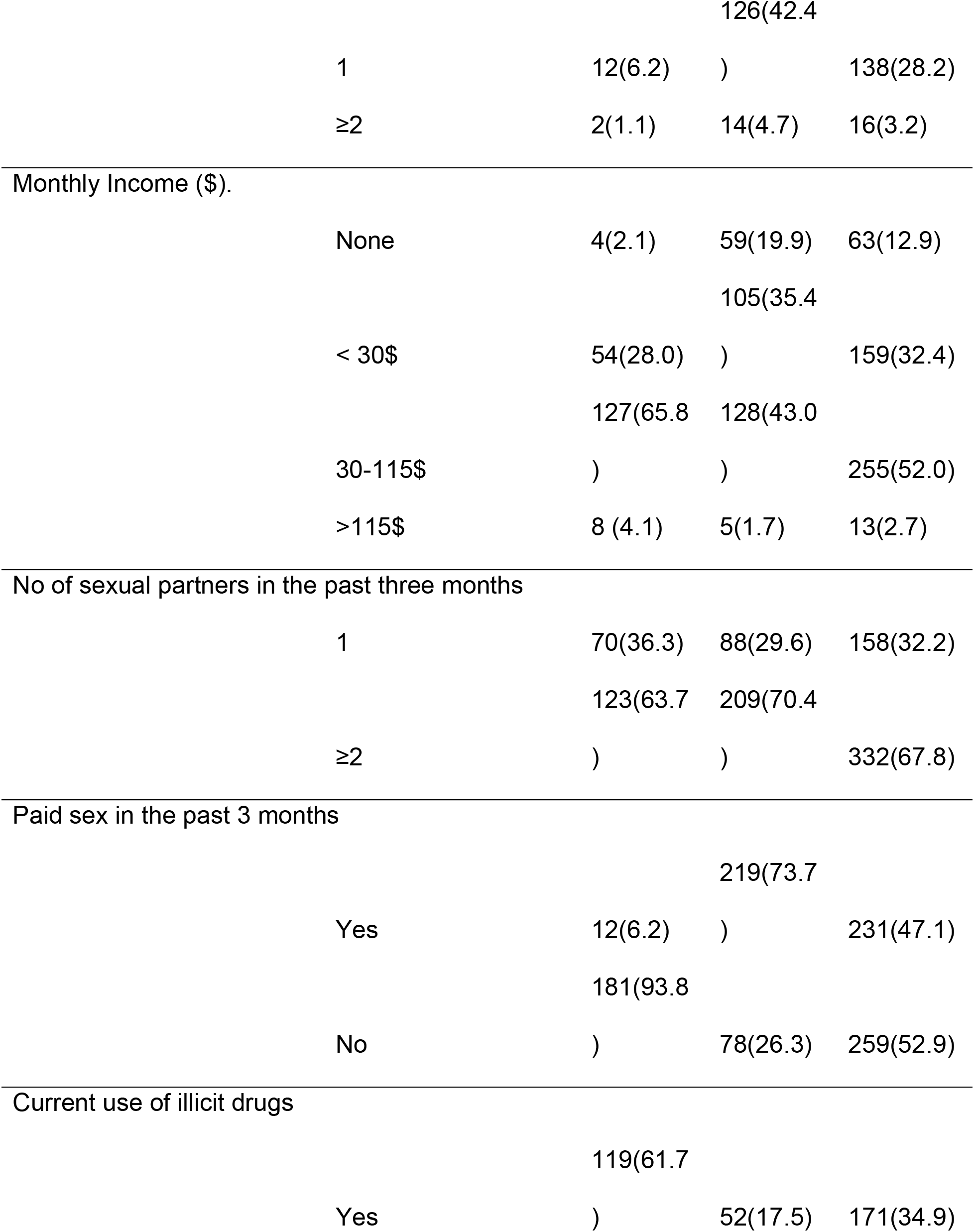
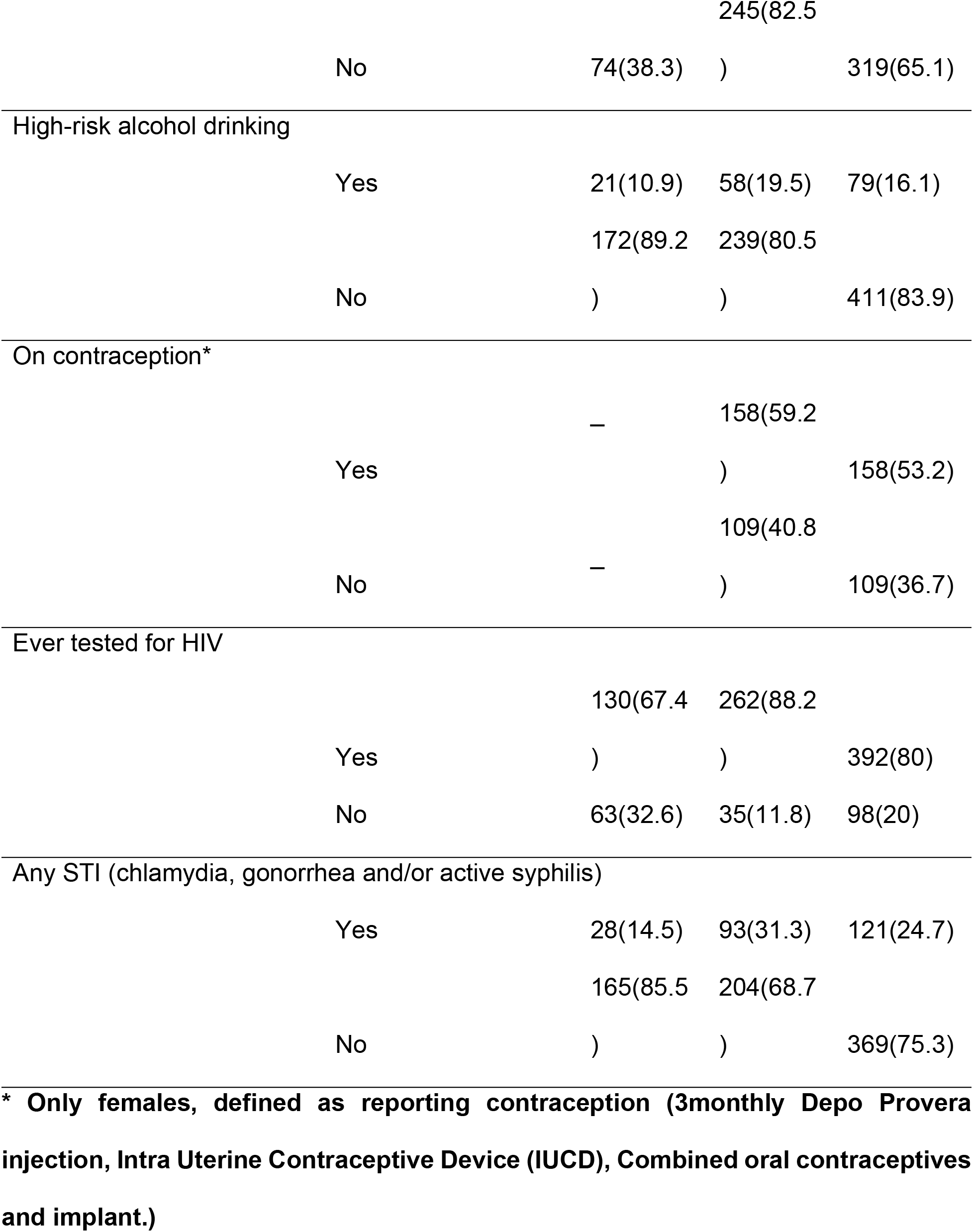
Baseline Characteristics of Adolescents Enrolled from Kampala Slums, Uganda.

### 3.2. Illicit drug and high-risk alcohol use among adolescents

At enrollment, 224 participants had ever used illicit drugs of whom 141 (62.9%) were male. Current Illicit drug users (past 3 months) were 171 (34.9%) with the proportion of those using Illicit drugs among males significantly higher than females (69.6% vs 30.4%, p<0.001), and 86 participants (50.3%) of current drug users reported using drugs daily (76.7% male’s vs 23.3% females), 68 (39.7%) reported using drugs once a week (64.7% males’ vs 35.3% females) while the rest used drugs less frequently. The most commonly used drugs for both males and females were marijuana (49.7%) and Khat (15.8%). Reasons given by males for taking illicit drugs were: to forget their problems (31.9%), to feel good (24.4%), peer pressure (11.8%), to get courage so they could do their work (3.4%) and other reasons (28.6%). Reasons given by females were peer pressure (28.9%), to feel good (23.1%), to get courage to engage in sex work (19.2%), and other reasons (28.9%).

Of the 490 participants, 79 (16.1%) were screened as high-risk alcohol users. And the proportion of high-risk alcohol use was 58 (73.4) among females Vs 21(26.6) males. At the unadjusted analysis, high-risk alcohol use was more likely among those who identified their main job as sex work, those with ≥10 paying sexual partners in the last 3 months, those who experienced any violence (Physical, sexual and emotional) from sexual partners in the last three months prior to the interview. The median (Interquartile range, IQR) of the audit score among high risk alcohol users was 20 (17–23) while among the non-high risk alcohol users was 1(0–6).

### 3.3. Baseline characteristics associated with illicit drug

At adjusted analysis, baseline characteristics associated with illicit drug use in the past 3 months were male gender (aOR 11.1; 95% CI 6.78-18.1) compared to females, being married (aOR 2.01; 95%CI 1.02-3.96) compared to being single and having 10 or more paying sexual partners (aOR 3.44; 95%CI 1.56-7.55) compared to reporting fewer than 10 (Table 2).

**Table 2:**
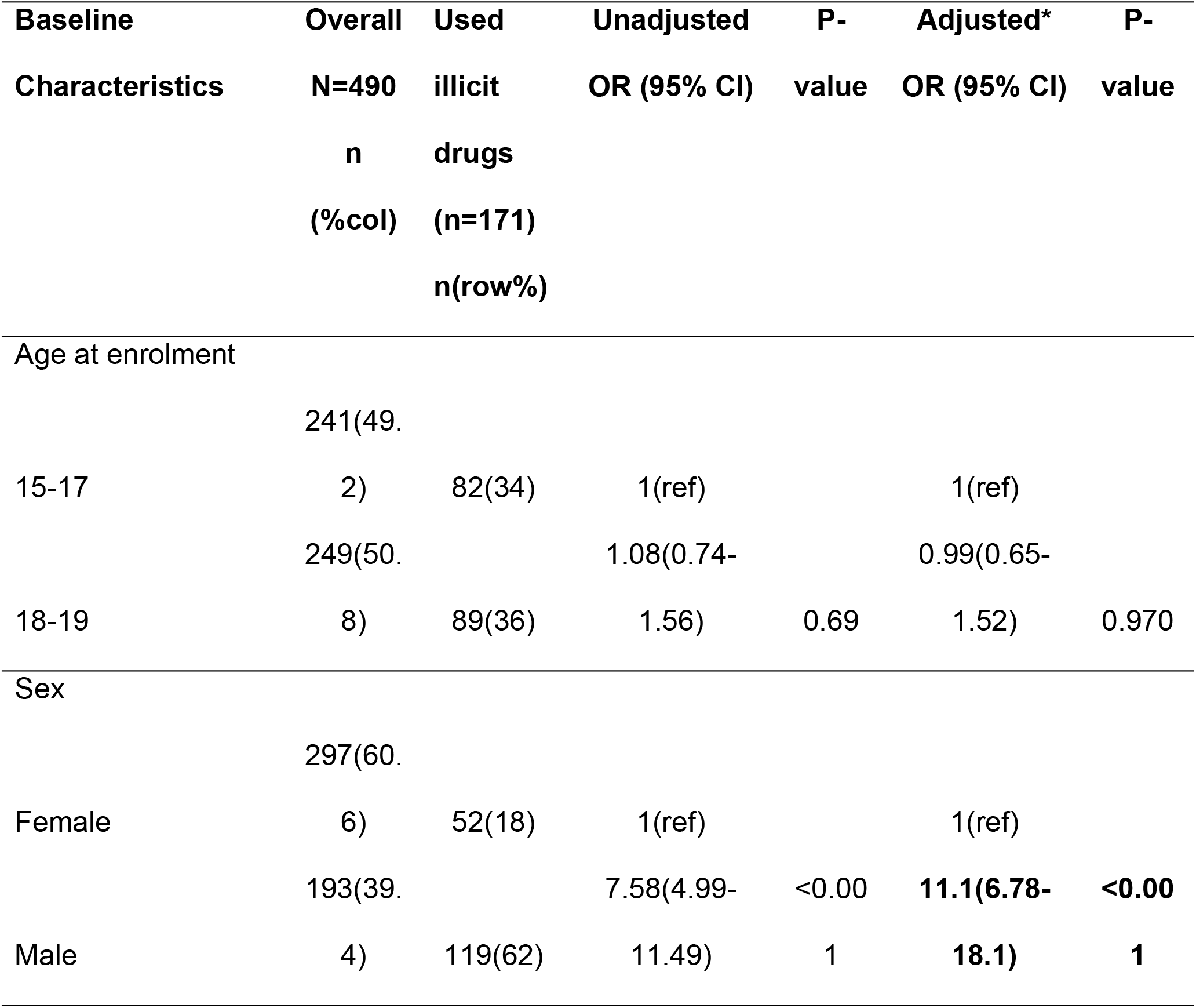
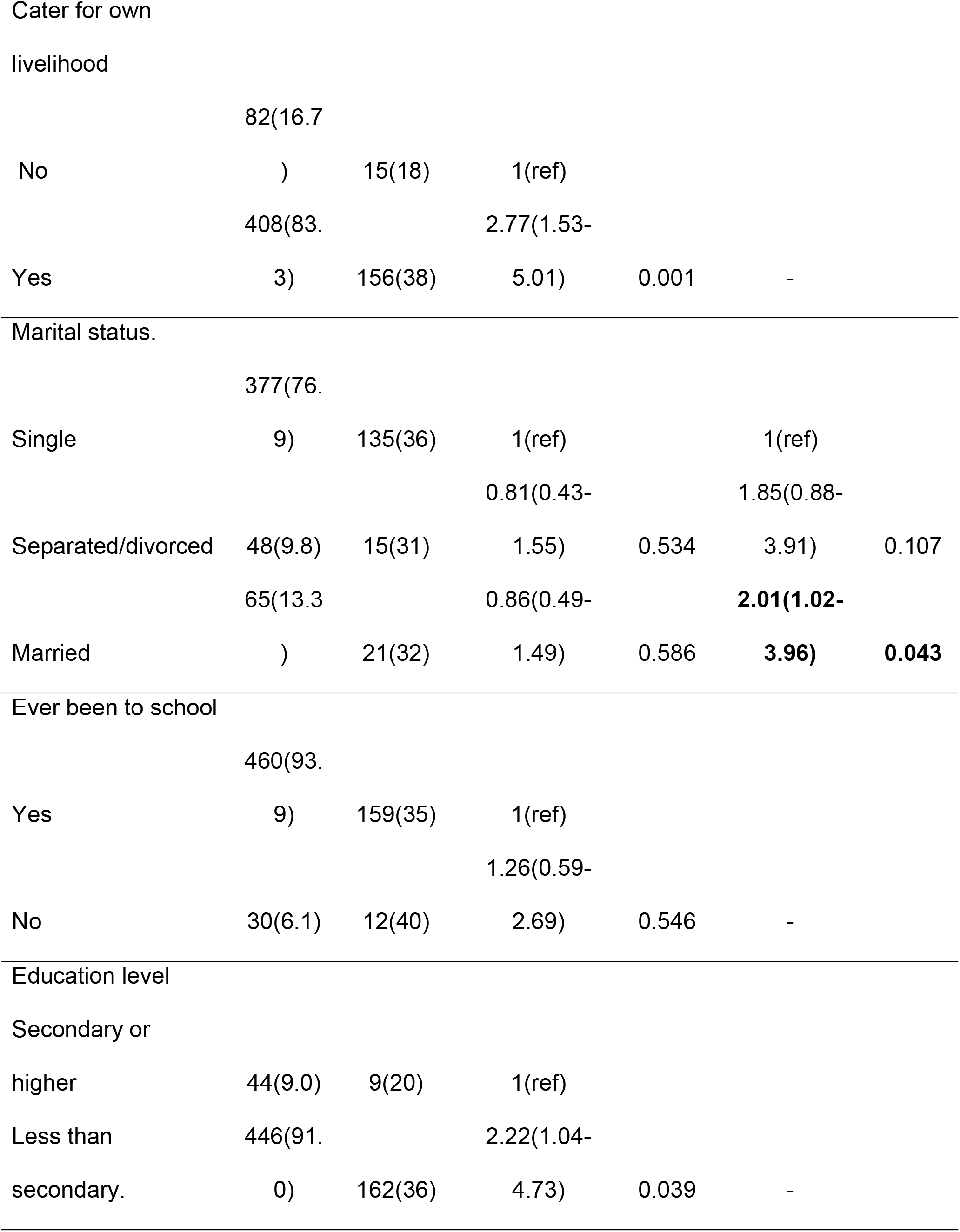
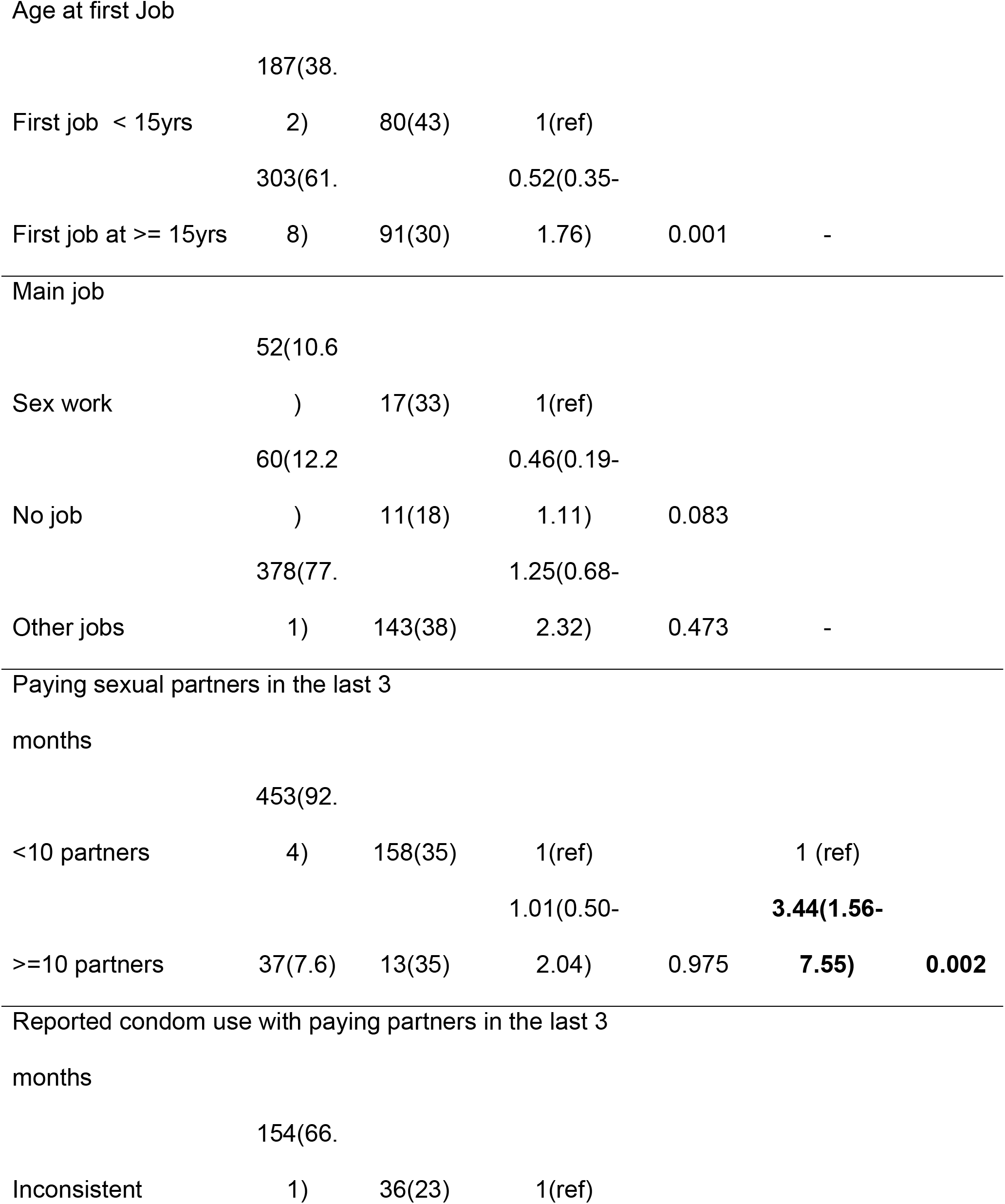
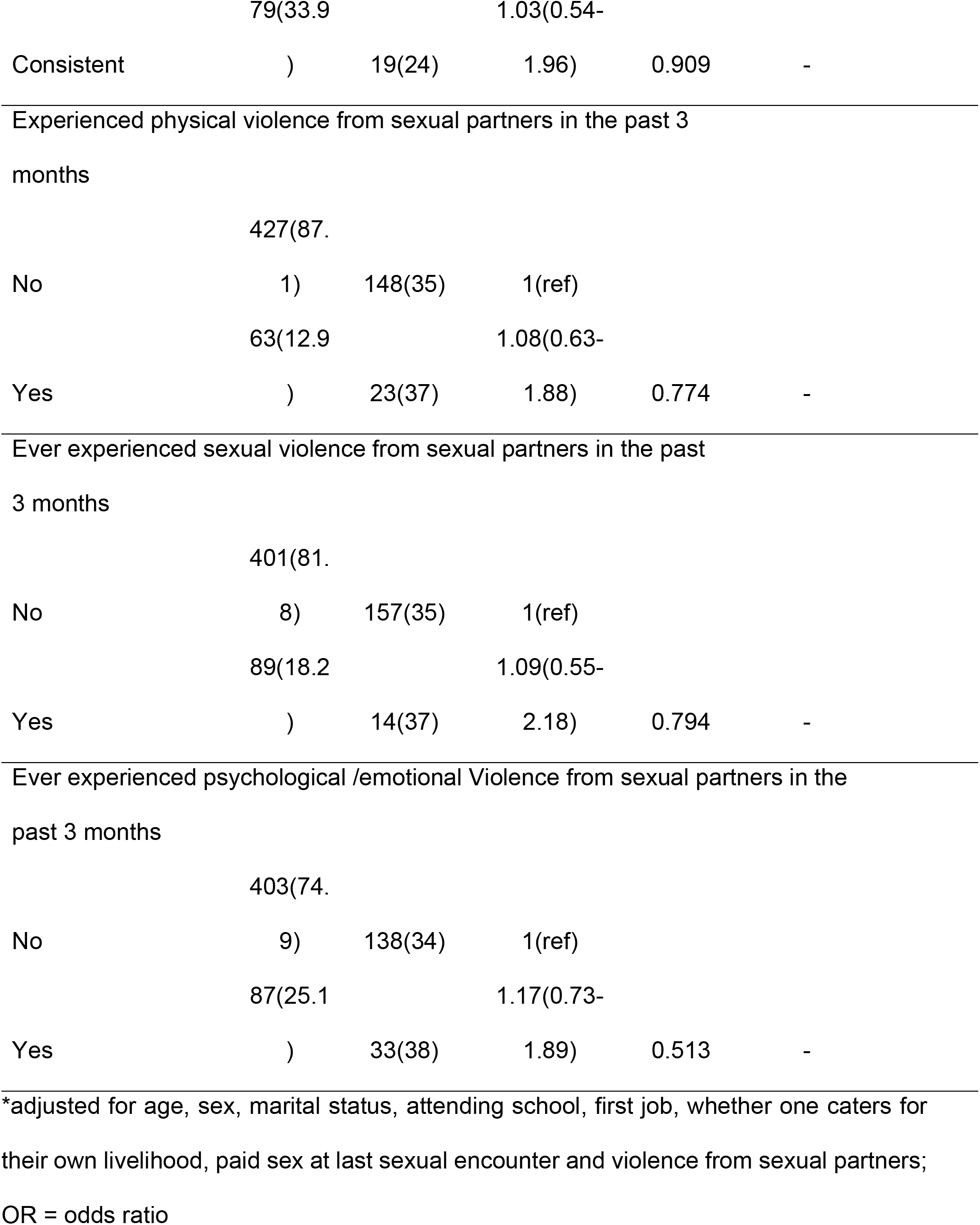
Association of participants’ baseline characteristics with illicit drug use in the past 3 months.

### 3.4. Baseline characteristics associated with high-risk alcohol use

At adjusted analysis, baseline characteristics associated with high-risk alcohol use were reporting sex work as a main job (aOR 4.22; 95%CI 1.49-11.9) compared to being unemployed and having experienced physical (aOR 2.02 95%CI 1.06-3.87) and or emotional violence (aOR 2.35 95%CI 1.32-418) from sexual partners, compared to those who did not report these respective types of IPV (Table 3).

**Table 3:**
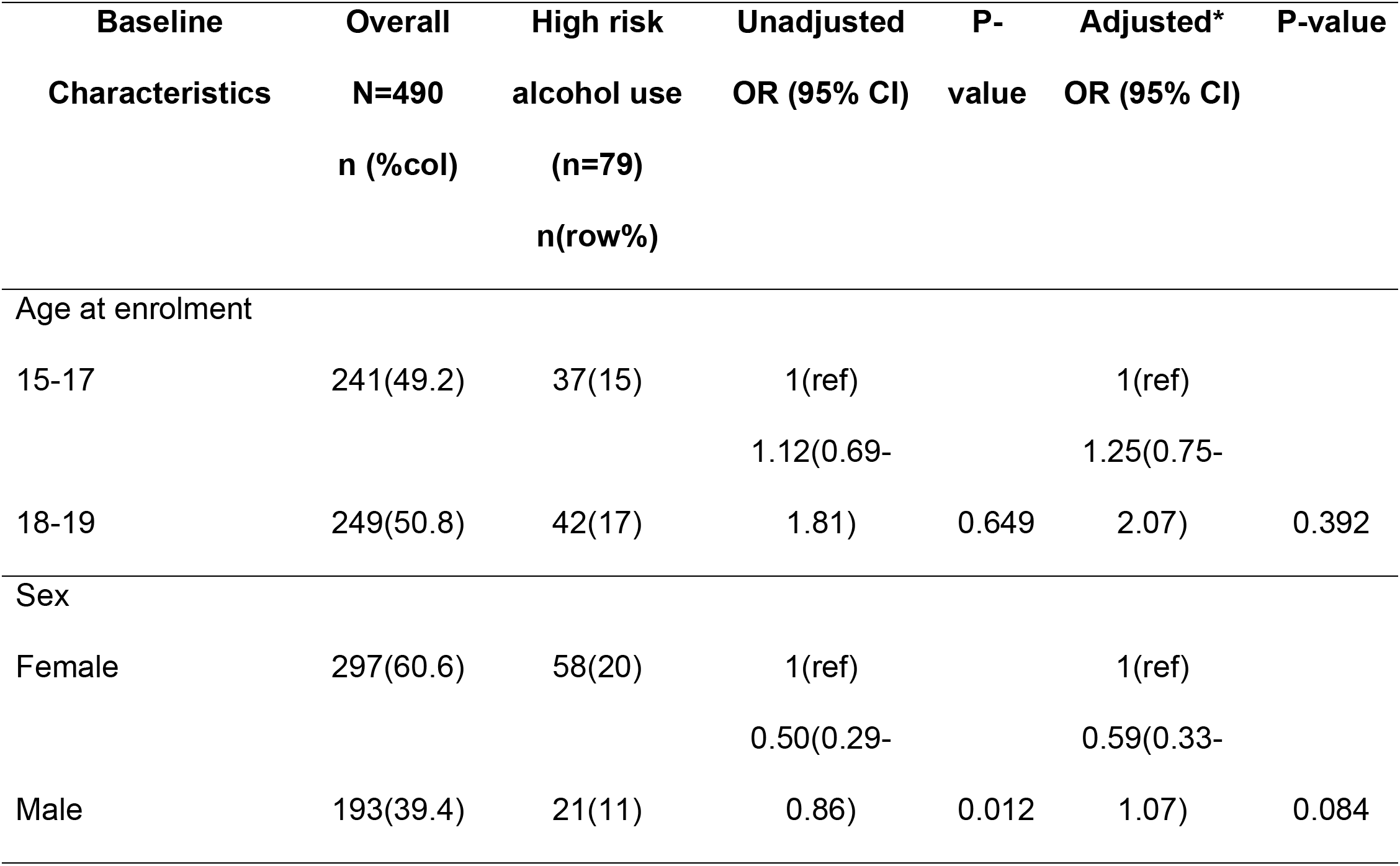
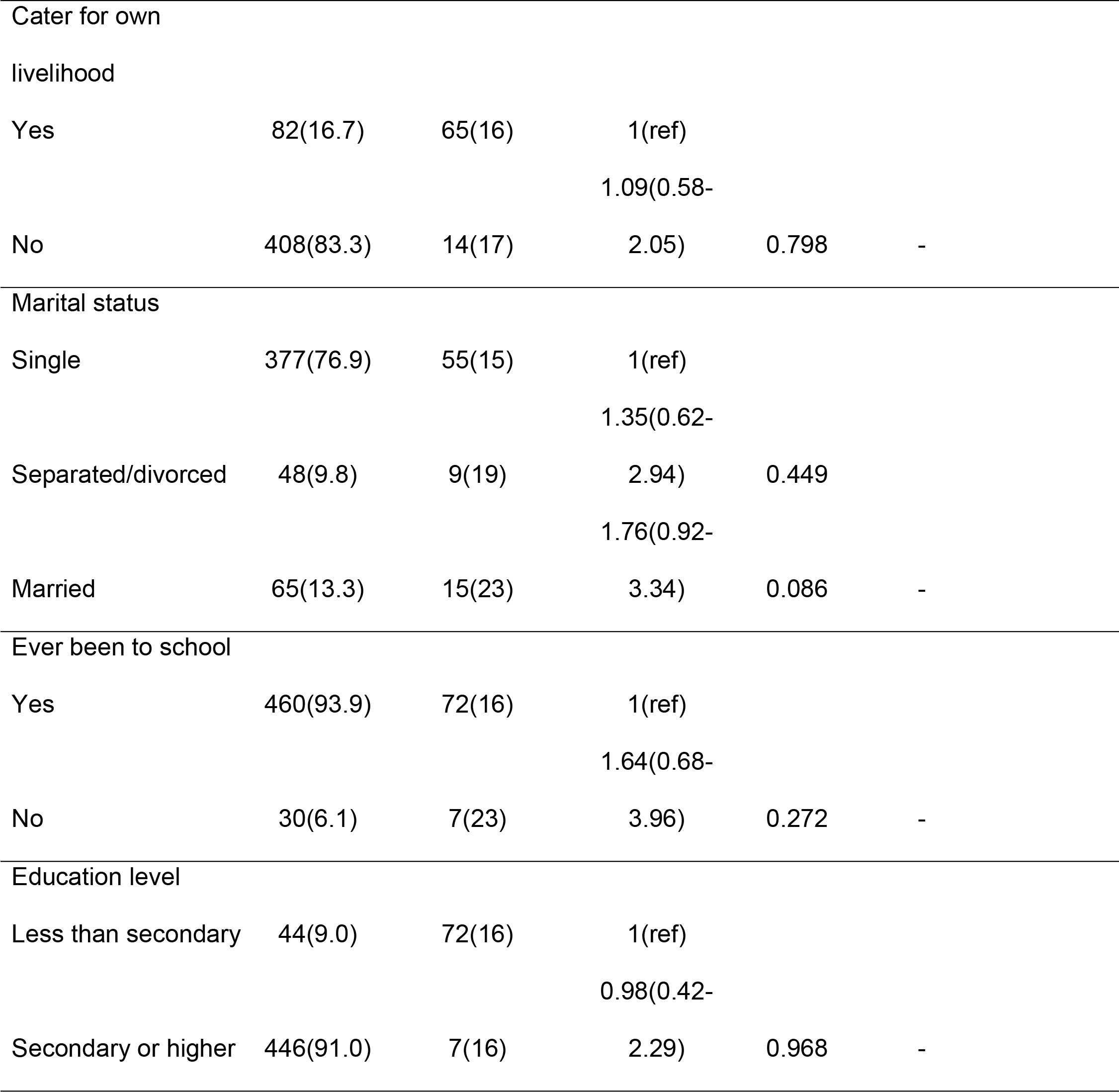
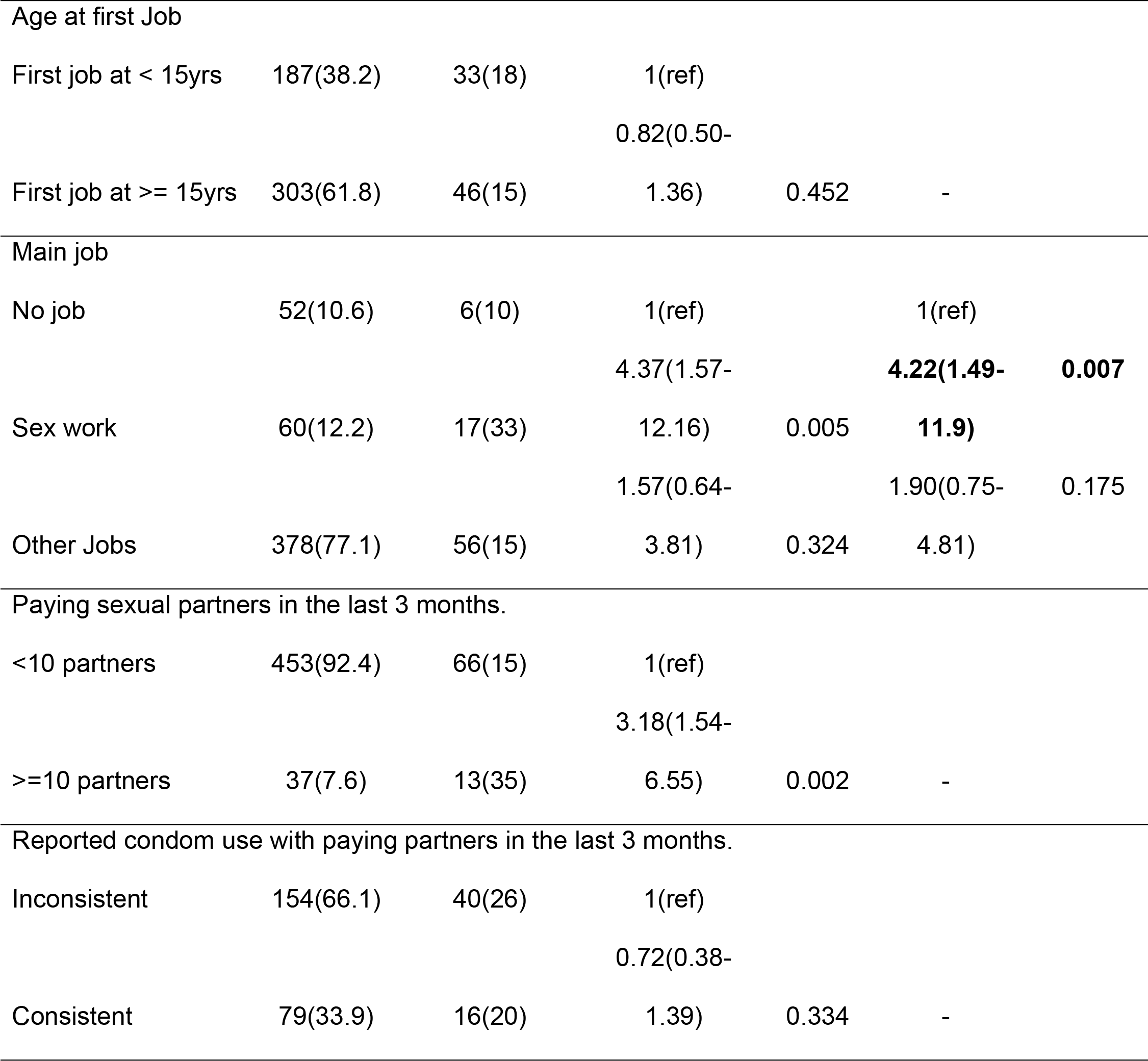
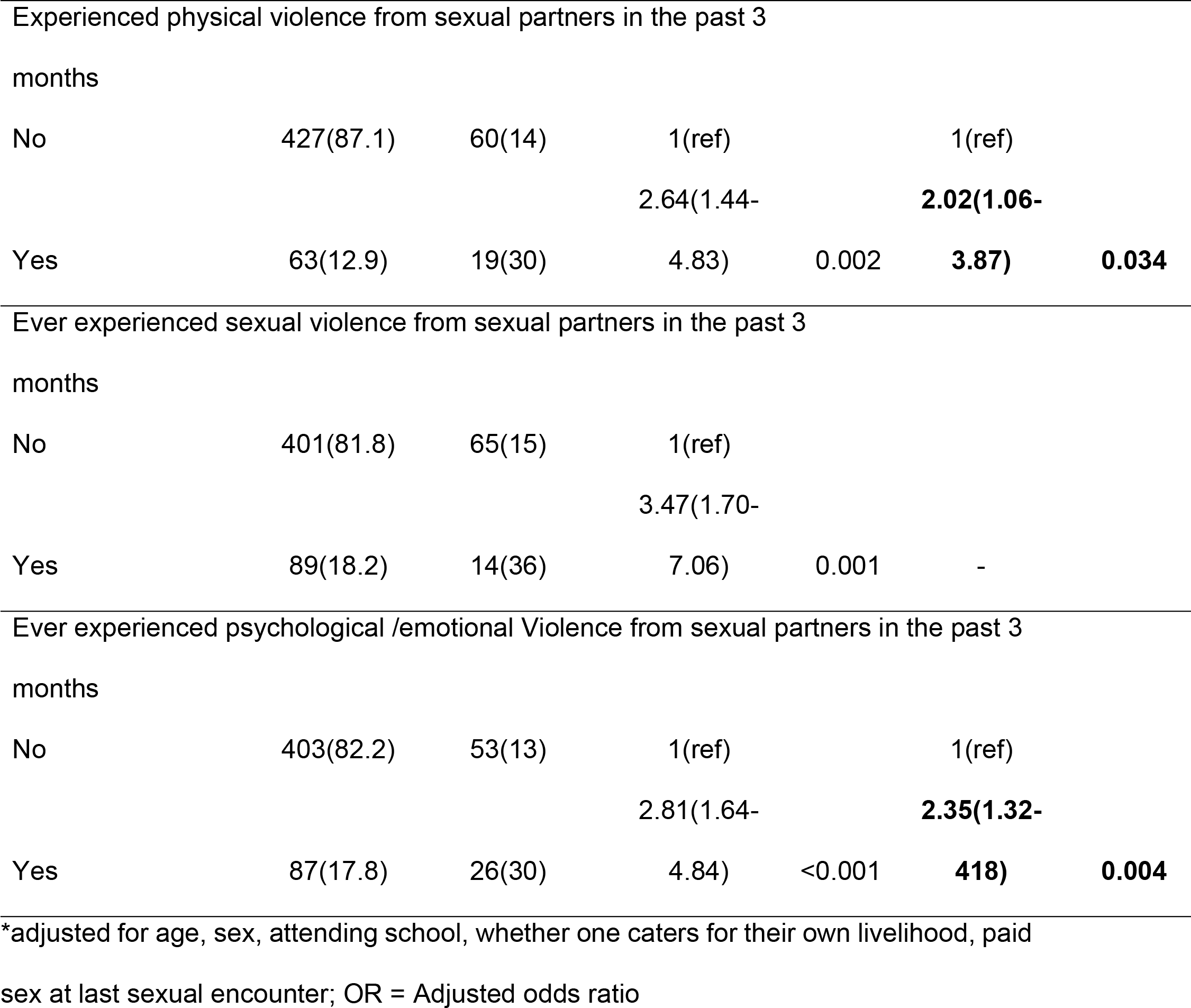
Association of participants’ baseline characteristics with high risk alcohol use.

## 4. Discussion

Our study found a high prevalence of illicit drug and high-risk alcohol use among 14-19-year old adolescents, and this is similar to other studies done among at-risk adolescents in similar vulnerable situations (15, 20). This is perhaps not surprising, given that the source GHWP clinic provides services to participants from Kampala slums that are characterized by a high prevalence of crime, illicit drug and alcohol use (25); this environment exposed adolescents to use these substances. However, our findings differ from a study done by Berhane et al in nine communities in SSA that reported very low prevalence of alcohol and illicit drug use among both school and non-school going adolescents 10-19-years (14), which the authors suggest may be attributed to under reporting. We found that adolescents who most abused drugs were taking care of themselves with no or under employment and this is in line with the recent warning from UNODC that increasing drug use rates are becoming more pronounced in countries with low levels of income (1).

Illicit drug use in our study was associated with being male and is consistent with similar studies done in Uganda, SSA and other parts of the world (10, 26, 27). Male participants in our cohort were married young, some had more than10 paying sexual partners in the last three months before enrolment and some experienced IPV from their sexual partners, they therefore likely resorted to use of drugs to mainly forget their problems caused by adolescent marriages, experiences of IPV from sexual partners (28, 29) and also to feel good. Indeed, we observed in this study that more than half of male participants who used drugs reported using drugs as a coping mechanism to mainly forget their problems and to feel good (21, 30). We further found that marijuana and khat were the most used drugs by adolescents (Figure1) which is similar to studies done in Uganda and SSA (10, 11, 21). These two drugs are locally available and cheap to purchase which makes it more accessible and affordable to adolescents with low levels of income compared to other drugs (10).

We found that illicit drug use was associated to being married which differs from another study done in Uganda in fishing communities that portrayed association of illicit drug use to being single (10), this was attributed to the fact that most of the married participants in this study got married young, and studies done in SSA show that adolescents living in informal settlements are more likely to be staying on their own, with friends or indulge in early marriages and thus lack formal parental control which expose them to riskier behaviors such as drug use, sex work, Intimate Partner Violence as observed in this study with subsequent high maternal and child mortality (28, 29, 31, 32)

Our findings also indicated that participants who reported engaging in sex work as their main job were 3-times more likely to be screened as high risk alcohol users compared to those who reported other main jobs, and having 10 or more paying partners was also associated with high risk alcohol use (8). Almost three quarters of our enrolled females reported engaging in paid sex in the past 3 months and most had low or no education. Sex work has been associated with alcohol use because women consume alcoholic drinks that clients buy for them (33), sex work sometimes occurs in bars and similar venues that sell alcohol (16, 33) and women report that taking alcohol gives them confidence to deal with many clients (20). Some also started sex work as their first job so took alcohol to cope with the stress of their job and other stressful issues linked to sex work e.g. discrimination, criminalization and also to cope with the cold (33, 34). The length of time involved in sex work also contributes to long-term alcohol use which can progress to high risk alcohol use over time (20, 33). Others likely abused alcohol due to peer influence around them given similar age group and involvement in similar high risk behavior such as sex work. Previous research has found that adolescents who perceived their friends as drug users, socialized with drug using peers and used alcohol for coping or fun were at increased risk of illicit drug, alcohol use and misuse (35). A much broader understanding of adolescent peers and their behavior may be one of the factors in curbing illicit and high-risk alcohol use among adolescents living in urban slums.

We further found that adolescents who experienced physical and emotional violence from sexual partners were more likely to be high risk alcohol users (20, 36). Most of our study participants engaged in paid sex, and it has been shown that when compared to older women, young women who engage in paid sex or sex work are more vulnerable to intimate partner violence from clients (37, 38). Younger women may also struggle with negotiation skills e.g., condom use, money paid per sex act, and this disadvantage may further expose them to acts of violence (34, 39). Given that sex work sometime takes place in venues that sell alcohol, IPV may also be perpetrated by disinhibited sexual partners/ clients who consume alcohol (40). For clients who chronically engage in IPV or engage in severe forms of partner violence, alcohol interventions may have a beneficial impact and the role of alcohol in the episodes of IPV was emphasized during counseling (36). Many of these issues correlate to social economic status (SES) where women with a partner who used alcohol and was of a lower SES had higher odds of experiencing IPV compared to women with partners of higher SES (40).

### 4.1. Limitations

Our study used a cross-sectional study design which limits our ability to infer causality in the associations seen between the dependent and independent outcomes. Our study was done among a unique population of adolescents that also included emancipated minors and findings may not be generalizable to adolescents in different settings and those who need parental/ guardian consent.

## 5. Conclusion

Illicit drug and high risk alcohol use were prevalent among adolescents from urban slums in Kampala. Male adolescents and those who report vulnerabilities like IPV, multiple sexual partnerships and sex work have a higher prevalence of substance use. Comprehensive interventions that reduce illicit drug and high-risk alcohol use are needed among adolescents living in urban slums to prevent negative long-term health consequences that may persist into adulthood.

## Competing Interests

The authors have no conflict of interest to declare

## Availability of data and materials

The datasets used and analyzed during the current study are available from the corresponding author.

## Funding Source

The project is part of the EDCTP2 program supported by the European Union (TMA2016CDF-1574). The views and opinions of authors expressed herein do not necessarily state or reflect those of EDCTP.

## Data Availability

The datasets used and analyzed during the current study are available from the corresponding author and can be shared with the journal when required.

## Acknowledgement

We wish to acknowledge the support from the University of California, san Francisco’s international traineeships in AIDS prevention studies. (ITAPS) U.S NIMH, R25MH123256. We acknowledge study participants, community team and the research team for their dedication to the study.

## Supporting Information

**S1 Fig. Percentages of illicit drugs used by adolescents.**

## Notes

### Competing Interest Statement

The authors have declared no competing interest.

